# A Plasma-based Deep Proteomic Platform for Early-Stage Breast Cancer Detection

**DOI:** 10.1101/2025.09.22.25336353

**Authors:** Alec Horrmann, Yash Travadi, Kevin Mallery, Grant Schaap, Kaylee Judith Kamalanathan, Nathaniel R. Bristow, Catalina Galeano-Garces, Song Yi Bae, Harrison Ball, Alexa R. Hesch, Sarah Pederson, Badrinath R. Konety, Yuliya Olimpiadi, Justin M. Drake

## Abstract

Despite the widespread use of mammography as the standard of care for breast cancer screening, its accuracy remains limited for select patient populations, such as women with high breast density. Liquid biopsy-based tests offer an accessible complement to conventional screening methods. Here, we conducted a case-control study to develop a plasma-based protein classifier to distinguish between early-stage breast cancer patients and healthy individuals. A total of 335 women, comprising 116 patients with newly diagnosed, treatment naïve breast cancer (Stage 0-2) and 219 healthy controls, had plasma samples collected and processed in a blinded manner using a sample preparation method coupled with semi-quantitative, label-free mass spectrometry (MS)-based analysis. The median number of proteins detected per patient across breast cancer and healthy individuals was 6,991 and 6,818, respectively. A machine learning-based classifier was trained and validated on patient proteome profiles using a leave-one-out cross-validation (LOOCV) approach to identify breast cancer patients. The classifier achieved an AUC of 0.96 (95% CI: 0.93-0.97), with a sensitivity of 86.2% (95% CI: 78.8-91.3%) and a specificity of 90.4% (95% CI: 85.8-93.6%). In breast cancer patients, the classifier retained >85% sensitivity regardless of breast density (low density: 87.2%, high density: 90.2%) at 90% specificity. Our workflow demonstrates the potential of plasma proteomics as a potent diagnostic tool in early-stage breast cancer screening.

## Introduction

Breast cancer patient survival improves drastically when detected early, with many localized cases being curative with surgical intervention [1]. Mammography remains the gold standard in breast cancer detection, with biennial screenings recommended by the U.S. Preventive Services Task Force (USPSTF) for women starting at age 40. However, inconsistencies in mammographic detection, particularly in women with dense breast tissue, can lead to inaccurate diagnosis [2]. The American College of Radiology utilizes the Breast Imaging Reporting and Data System (BI-RADS) classification system to grade breast density on a scale from A (lowest density, mostly fat) to D (highest density, mostly glandular tissue). Approximately 50% of women above the age of 40 have dense breast tissue (defined here as BI-RADS C or D), and women with dense breasts have a higher risk of developing breast cancer [3]. While alternative imaging methods, such as MRI, digital breast tomosynthesis (or 3D mammography), contrast enhanced mammography (CEM), and ultrasonography are available and recommended for some high risk women such as those with a BRCA mutation, there is no clear consensus on guidelines for their use in patients with high breast tissue density who do not meet other “high risk” criteria [4], [5]. Consequently, these tests are often not covered by insurance for the general population, leading to broad inaccessibility and high out-of-pocket costs. Hence, there is a strong need for diagnostic tests capable of detecting early-stage breast cancer that are both sensitive and accessible to the public.

Liquid biopsies offer a promising complement to conventional screening methods, with the potential to vastly improve the accessibility of early cancer detection. These tests screen biological fluids, such as blood, sampled through standard medical procedures for various cancer analytes, including cell free DNA (cfDNA), circulating tumor DNA (ctDNA), microRNAs, orphan non-coding RNAs (oncRNAs), and proteins [6]. There are currently no FDA approved liquid biopsy tests to detect early-stage breast cancer. Galleri, a multi-cancer detection assay developed by GRAIL, is a commercially available blood test that screens patients using targeted methylation analysis of ctDNA [7]. While the specificity of this assay is high (>99%), the sensitivity for early-stage breast cancer detection is low (2.6%-47.5% for stage 1 and 2 patients, respectively) [8], likely due to the low shedding rate of ctDNA in early-stage cancers [9]. Several protein biomarkers have been approved by the FDA for treatment monitoring in breast cancer, including CA 15-3, CA 27-29, and carcinoembryonic antigen (CEA). However, they lack sufficient sensitivity and specificity for early detection [10]. More sensitive technologies will be required to explore the full proteomic landscape in early-stage breast cancer.

The plasma proteome is highly complex, estimated to contain over 10,000 unique proteins ranging more than 10 orders of magnitude in abundance [11]. A subset of these targets is heavily overrepresented in plasma, limiting the detection of low abundance proteins [12]. Innovations in plasma sample processing, such as the depletion of high abundance proteins and sample multiplexing, have revolutionized our ability to detect clinically relevant proteins. Liquid chromatography-tandem mass spectrometry (LC-MS/MS, hereafter referred to as MS in this study) is an unbiased method of protein quantification capable of identifying a wide range of proteins [13], [14]. These techniques often generate complex datasets, comprising thousands of peptide fragments correlated to hundreds of proteins or more. Consequently, studies aimed at disease detection tend to focus on select panels of differentially expressed proteins, which may not capture the intricacies of the proteomic landscape and lead to diagnostic inaccuracies [15]. Recent studies have utilized machine learning methods to help overcome these challenges, correlating complex proteome profiles to disease status [16], [17], [18].

Here, we present a liquid biopsy-based proteomics workflow for early-stage breast cancer detection aimed at overcoming the current limitations of low specificity in this setting while achieving high sensitivity. Astrin Bioscience’s sample preparation workflow combines protein enrichment with semi-quantitative, label-free MS-based analysis to enhance the dynamic range of proteins detected within each sample. Raw MS profiles are processed using a breast cancer-focused, empirical spectral library to quantify greater than 6,500 proteins per patient on average. We trained a protein-based machine learning classifier to identify patients with early-stage breast cancer. This unbiased approach to protein analysis incorporates a large proportion of the detected proteome within each patient, expanding the total number of proteins used for diagnostic assessment. Collectively, this study demonstrates a streamlined, high-throughput workflow for early-stage breast cancer detection while addressing several key limitations with modern proteome analysis.

## Materials and Methods

### Sample Acquisition and Processing

This was a case-control, observational study. Early-stage breast cancer patients were recruited as part of a local study at Allina Health Cancer Institute (Minneapolis, MN), and additional samples were sourced from the contract research organization (CRO) iSpecimen (Woburn, MA). Women over the age of 18 were eligible for inclusion as cases in this study if they were 1) treatment naïve with 2) no prior history of cancer and 3) had a confirmed diagnosis of stage 0-2 breast cancer based on histopathological analysis. All breast cancer cases had either a screening or diagnostic mammogram prior to study enrollment. Healthy women donors were recruited at Astrin Biosciences (Saint Paul, MN) and sourced from iSpecimen and ZenBio (Durham, NC). Healthy women donors aged 18-74 were self-reported healthy and considered eligible if they had 1) no prior history of cancer, 2) had no first-degree relatives with a history of breast cancer, and 3) were not pregnant at time of enrollment. While a screening mammogram was not required for healthy controls in this study, women over the age of 35 had to have had a negative mammogram within one year of enrollment to be included. A higher proportion of younger, healthy individuals were selected for method development because patients under the age of 35 without a family history of breast cancer have less than a 0.3% chance of developing the disease within the next five years, increasing our confidence they are truly negative. For patients with either a screening or diagnostic mammogram, breast density was assessed during imaging and categorized using the American College of Radiology’s BI-RADS classification system. Within this study, women were further classified as having either “low density” breasts (BI-RADS A or B) or “high density” breasts (BI-RADS C or D). The studies were approved by the WCG Institutional Review Board and all participants provided written informed consent.

Blood samples were drawn from breast cancer cases at a follow-up appointment following biopsy confirmation but prior to any treatment. For both cases and controls, whole blood was collected into EDTA tubes and shipped overnight to Astrin Biosciences for processing. The status of each sample was randomized and blinded to laboratory personnel. Blood samples were double spun for plasma isolation, and the supernatant was transferred to a fresh conical each time before being aliquoted and stored at-80°C. Previously un-thawed plasma aliquots were then loaded into a Microlab Prep (Hamilton) robot liquid handler along with process and digestion controls, and positive and negative controls comprising pooled cancer patient and healthy plasma, respectively. Proprietary functionalized superparamagnetic bead solutions were added to each sample to allow for preferential protein capture while leaving behind contaminating high abundance proteins. In parallel to protein capture, incubation, wash, and lysis plates were created. These optimized buffers serve to assist in depleting weakly and non-specifically adhered proteins and to fully denature, reduce, and alkylate the proteins. The deep-well plate along with wash and lysis plates were transferred to a Kingfisher Flex (Thermo Fisher) that performed a series of gentle washes followed by heated lysis on a heated shaker. Using the Microlab Prep, the samples were transferred to a 96 well LoBinding plate (Eppendorf, 0030129512) for all subsequent steps. A protein recapture buffer was then added to promote optimal protein recovery. Following a series of wash buffer exchanges, the samples were transferred into a digestion buffer (pH 8.0) and digested with Lys-C and Trypsin overnight. The next morning, the beads were removed, and the peptide-containing supernatant was transferred and dried down on a vacuum concentrator (Labconco).

### Peptide Quantification and Mass Spectrometry Parameters

Both breast cancer cases and healthy controls were randomized within sample preparation batches prior to analysis. Peptides were resuspended and quantified using a quantitative fluorometric peptide assay ( Thermo Fisher, 23290) to normalize peptide quantities prior to loading onto EvoTips (EvoSep, EV2011). EvoTips were prepared as directed by the manufacturer with the exception that all volumes used were doubled. Samples were then analyzed on an Orbitrap Astral (Thermo Fisher) mass spectrometer coupled to an EvoSep One (EvoSep) running a whisper zoom 40 SPD method with a 15 cm Ion Opticks column (AUR3-15075C18-XT) and set to 1,800 V. The Astral was run with default parameters from the Label Free Quantification using Data Independent Acquisition (LFQ DIA) preset with 3 Th windows. Additionally, a commercially available protein digest was run sequentially within each sample batch to ensure mass spectrometer performance as well as automated instrument mass calibrations with every run.

### Mass Spectrometry Data Analysis

A empirical spectral library was generated using a subset of patient samples. All results were generated from DIA-NN (2.1.0) with the following settings: Maximum mass accuracy fixed to 11 and 10 ppm (MS1 and MS2 respectively), scan window of 7, trypsin/P, missed cleavages 1, peptide length between 7 and 30, carbamidomethylation as a fixed modification, up to one variable modification of methionine oxidation or N-terminal acetylation, and 1% FDR. A random subset of 100 patient samples was designated as “normalizing samples” for protein quantification. QuantUMS parameters were trained on these 100 samples and fixed for subsequent analysis [19]. The remaining samples were each processed in independent DIA-NN sessions alongside all 100 normalizing samples (i.e. total of 101 samples) to obtain the MaxLFQ-normalized quantifications [20].

The gene group-level abundance data from DIA-NN was preprocessed as follows. Razor peptides, peptides that are not unique to a single human protein, were assigned to the most likely corresponding gene, and abundances were aggregated by summing values across unique genes, treating missing values as zero during aggregation. Non-missing abundances were log-transformed, and any remaining missing values were imputed using the minimum observed abundance for each gene symbol minus one. Finally, the abundances were standardized by gene (zero mean and unit variance) to generate features suitable for machine learning classification models.

### Statistical Analysis to Develop a Cancer Classifier

We used logistic regression with L1 regularization (Lasso, least absolute shrinkage and selection operator) to classify breast cancer patients from healthy controls. We employed a leave-one-out cross-validation (LOOCV) strategy for model validation, i.e., for each sample a model was trained excluding the held-out sample, and prediction probabilities were obtained on that sample. Classification performance metrics were then computed by aggregating prediction outcomes across all samples. All data preprocessing steps, including missing value imputation and normalization, were performed within each LOOCV fold using only the training data and then applied to the held-out sample. Additionally, the Lasso penalization parameter was tuned within each LOOCV fold using a nested 5-fold cross-validation on the training set. All data analysis was performed using Python 3.12.4; the model was trained using the scikit-learn library (version 1.4.2).

## Results

### Proteomic characterization of the patient plasma

A total of 335 women were recruited for this study, including 116 newly diagnosed, treatment naïve Stage 0-2 breast cancer patients (stage 0: n = 30, I: n = 72, II: n = 14) and 219 healthy controls. The patient demographics are summarized in Table 1 and Table S1. The median age (Q1-Q3) in the cancer and non-cancer groups were 64 (52-70) and 32 (25-34), respectively. A higher proportion of younger healthy control individuals were specifically recruited to reduce the risk of false negatives and to improve the accuracy of our ML classifier during method development. Patients under the age of 35 without a family history of breast cancer have less than a 0.3% chance of developing the disease within the next 5 years. A total of 30.6% (n = 67/219) of the healthy controls included in this study were within the USPSTF recommended screening age (40-74) (Figure 1A). The racial demographics between the breast cancer and healthy control groups were roughly equivalent, with a larger proportion of white individuals in the breast cancer group relative to the healthy control group (84.5% vs 70.8%, respectively). Blood samples were de-identified and blinded before processing with Astrin’s sample preparation workflow (Figure 2A). All MS files were mapped using Astrin’s breast cancer-focused spectral library to identify and quantify proteome profiles.

**Figure 1.**
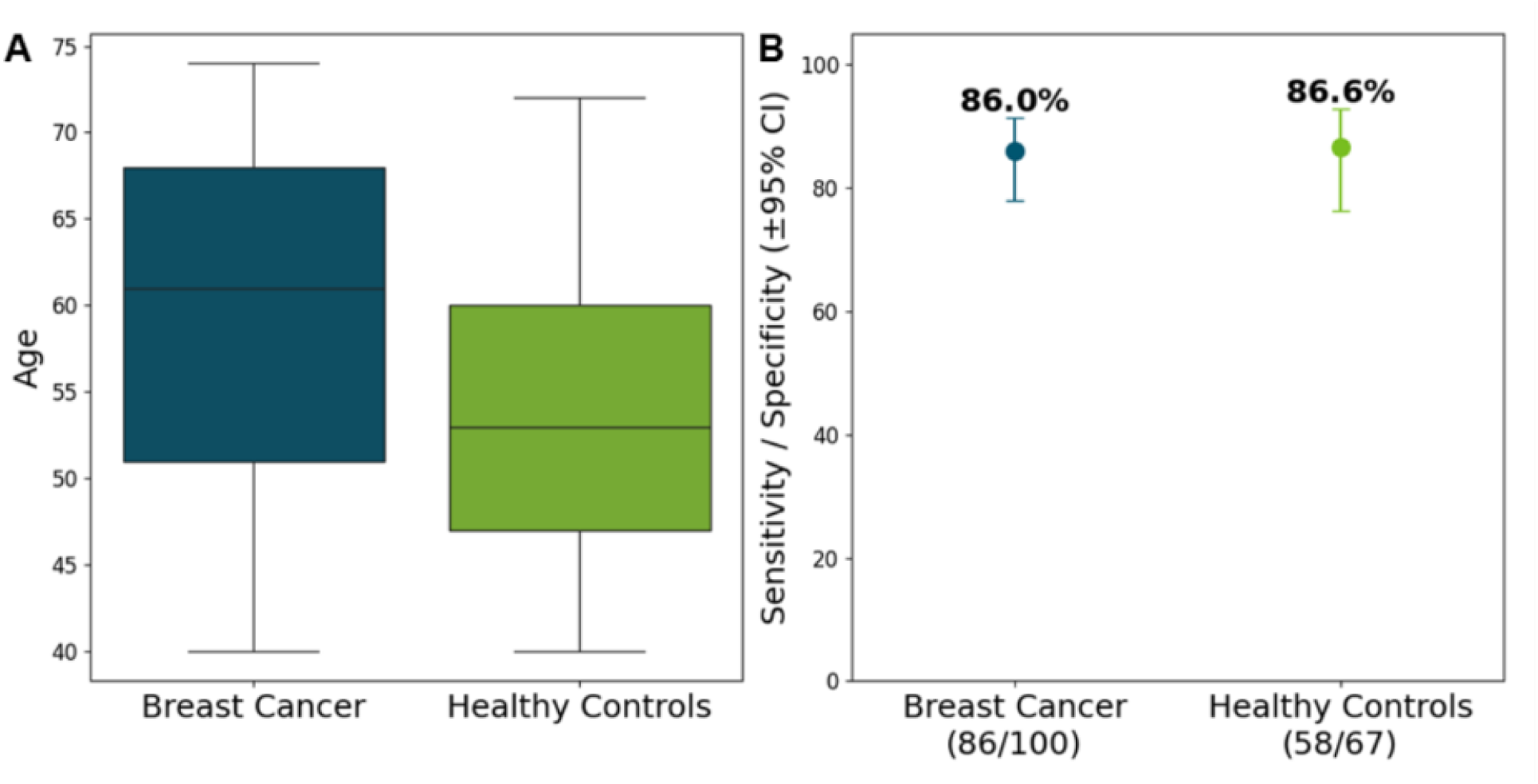
Evaluation of the protein-based classifier on individuals between the ages of 40-74. (A) Box plots of the distribution of ages across the breast cancer patients (n = 100) and healthy controls (n = 67). (B) Dot plot of the sensitivity and specificity of the classifier for the age-adjusted cohort.

**Figure 2.**
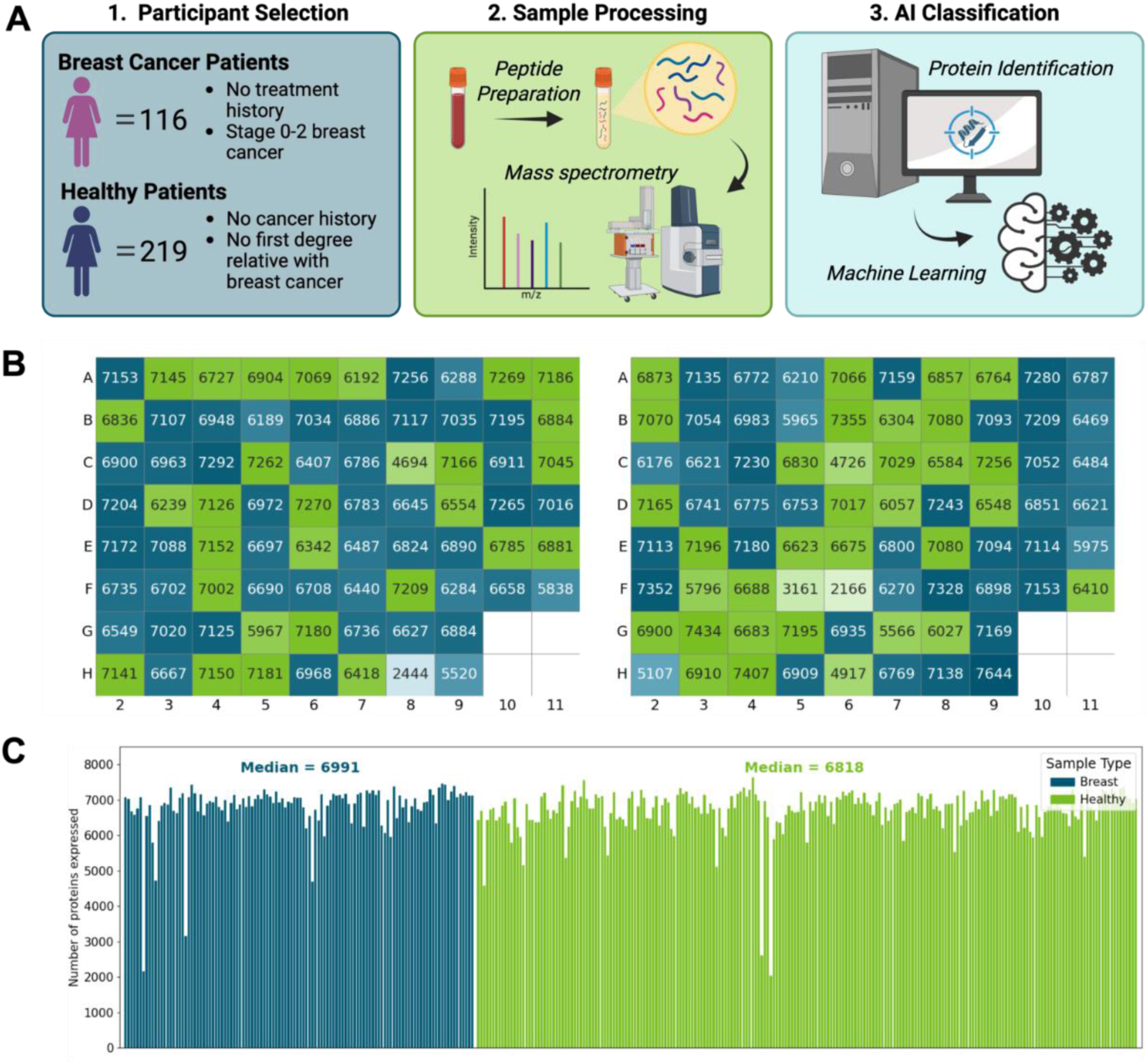
Overview of study design and summary of protein identification. (A). Blood was drawn from a total of 335 patients for this study, including 116 early-stage (Stage 0-2) breast cancer patients and 219 healthy control individuals. The breast cancer patients were newly diagnosed and treatment naïve, and the healthy control individuals were screened for previous and hereditary cancer history prior to enrollment. Samples were shipped overnight to Astrin Biosciences for processing and mass spectrometry analysis. The resulting patient proteome profiles were used to train a machine learning classifier to identify patients with early-stage breast cancer. (B) Well-plate schematic of the first two plates showing the number of proteins identified between breast cancer (blue) and healthy control individuals (green). The plate layout was randomized to minimize plate specific effects on the analysis. (C) Bar graph depicting the number of proteins identified per sample in breast cancer patients (blue) and healthy control individuals (green). Created in BioRender. Ball, H. (2025) https://BioRender.com/pbyhjdd

**Figure 3.**
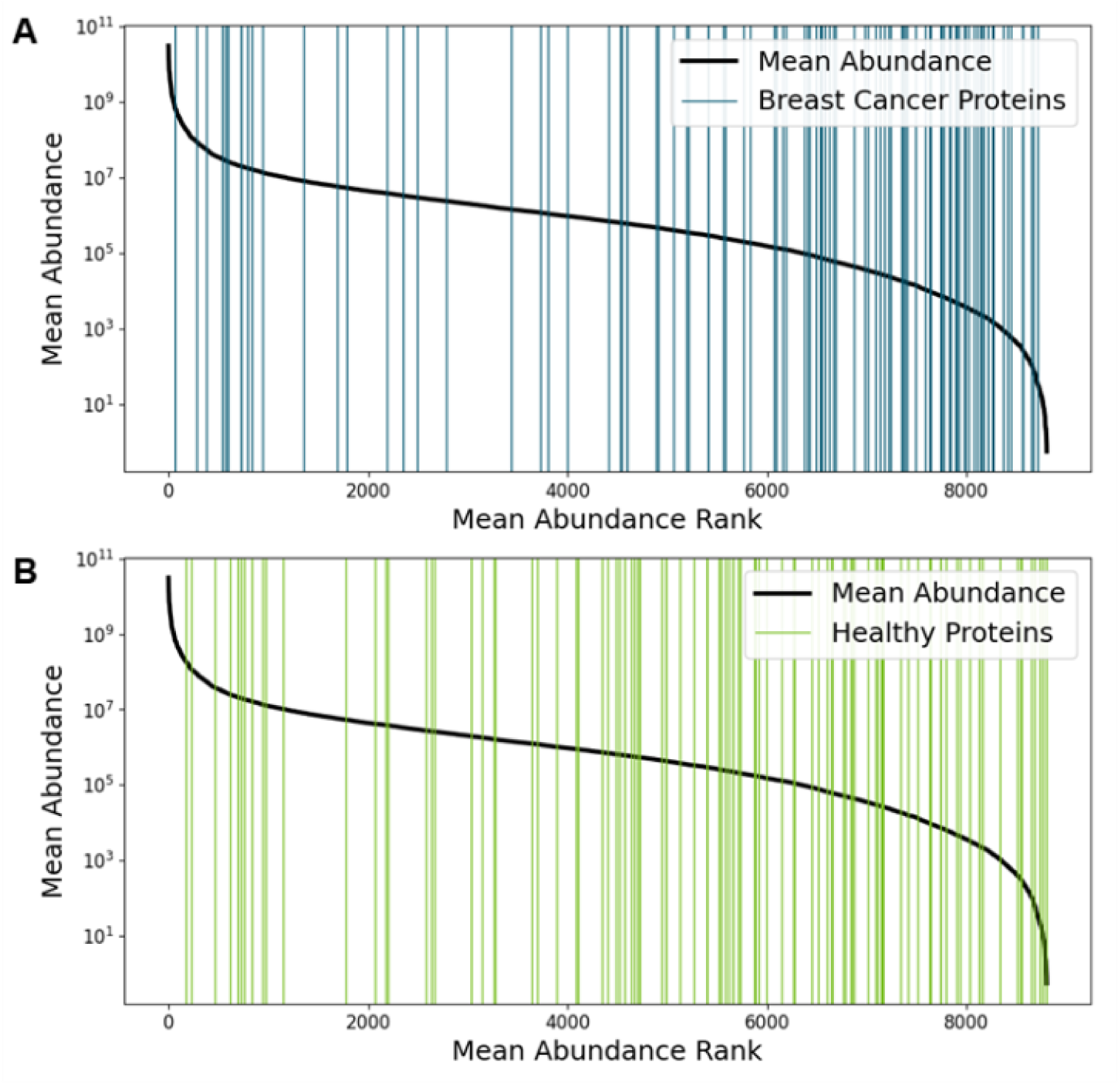
Rug plots denoting the mean abundance distribution of: (A) the top 100 breast cancer specific proteins selected by the classifier and (B) the top 100 healthy control specific proteins selected by the classifier. The increased concentration of cohort-specific proteins in the lower abundant regions highlights the importance of our deep proteomic profiling workflow in identifying markers differentially expressed across the two cohorts.

**Table 1.**
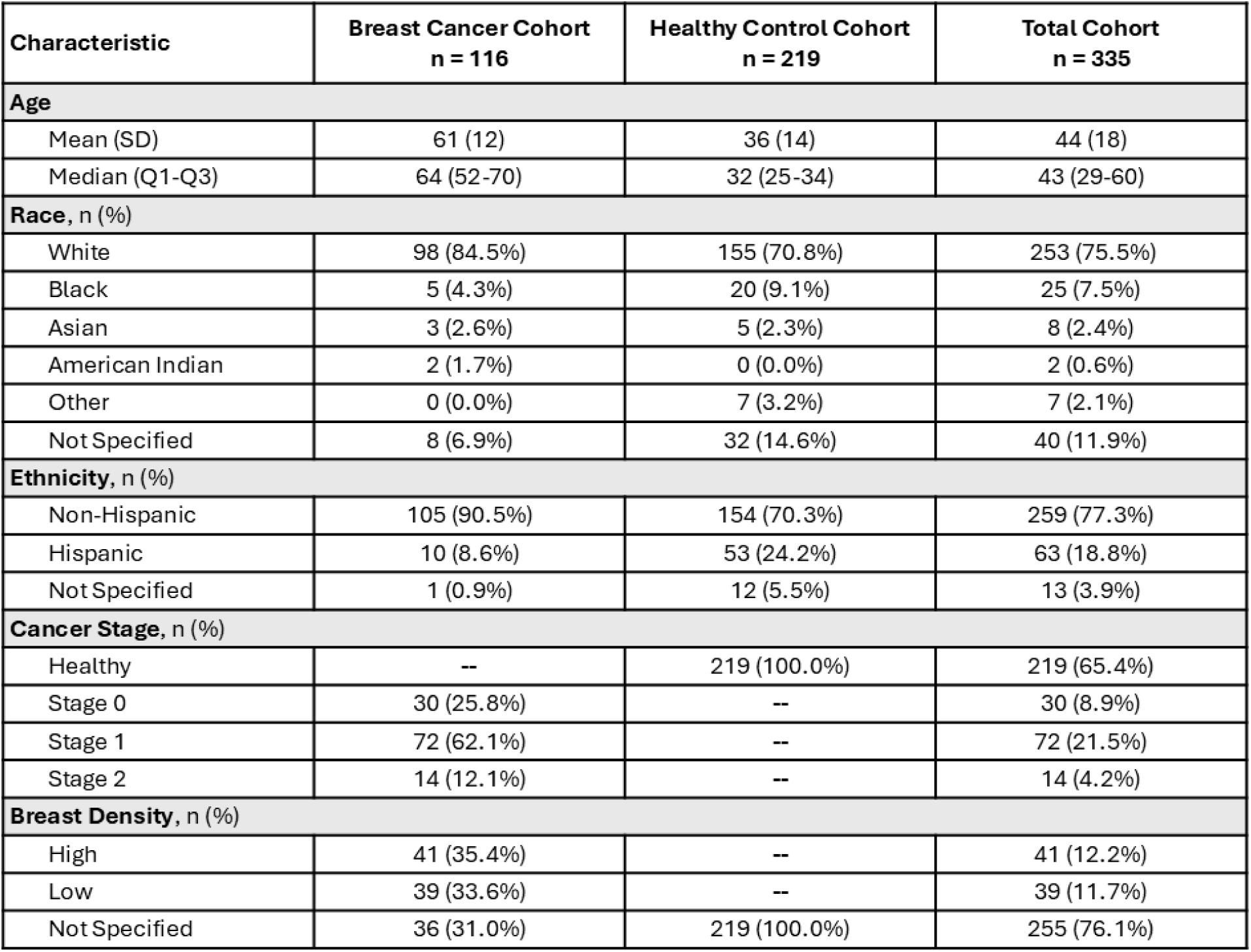
Summary of patient demographics between breast cancer cases and healthy controls.

The median (Q1-Q3) number of proteins detected between breast cancer cases and healthy controls were 6,991 (6,735-7,140) and 6,818 (6,499-7,100) proteins, respectively, with 9,030 unique proteins observed across both groups. Samples across the well plates were randomized to reduce plate specific effects on protein identification. Inspection of the number of proteins identified by well-plate showed no discernible patterns (Figure 2B) across groups, suggesting the absence of plate-based interactions. Most patients had more than 6,500 proteins detected per sample (78.2%, 262/335), demonstrating both the consistency of our sample preparation and the sensitivity of our MS method (Figure 2C). Quantitative assessment of protein intensities showed that we captured a dynamic range of protein abundances spanning over 8 orders of magnitude, highlighting the workflow’s potential to capture low abundance protein fractions (Figure S3).

### Protein-based classification of disease status

To aid in the interpretation of the quantified plasma proteins and identify patients with early-stage breast cancer, we trained a machine learning classifier on patient proteome profiles to distinguish between cancer and healthy individuals. The protein-based classifier achieved an AUC of 0.96 (95% Bootstrap CI: 0.93-0.97) in differentiating between early-stage breast cancer patients and healthy individuals. A Receiver operating curve (ROC) showed strong discriminative performance, demonstrating the model’s accuracy for early-stage breast cancer prediction (Figure 4A). Across all patients, the model achieved a sensitivity of 86.2% (95% Wilson CI: 78.8-91.3%) with a specificity of 90.4% (95% Wilson CI: 85.8-93.6%) (Figure 4B). To elucidate any age-related effects within the classifier, we also evaluated the model only on individuals within the recommended screening age for breast cancer (n = 167). The differences in the sensitivity and specificity between all patients and the age-adjusted patients were not statistically significant, with only a small decrease in specificity observed (sensitivity: 86.2% vs 86.0% (Two-sided Z-test p-value: 0.97); specificity: 90.4% vs 86.6% (Two-sided Z-test p-value: 0.37)) (Figure 1B).

**Figure 4.**
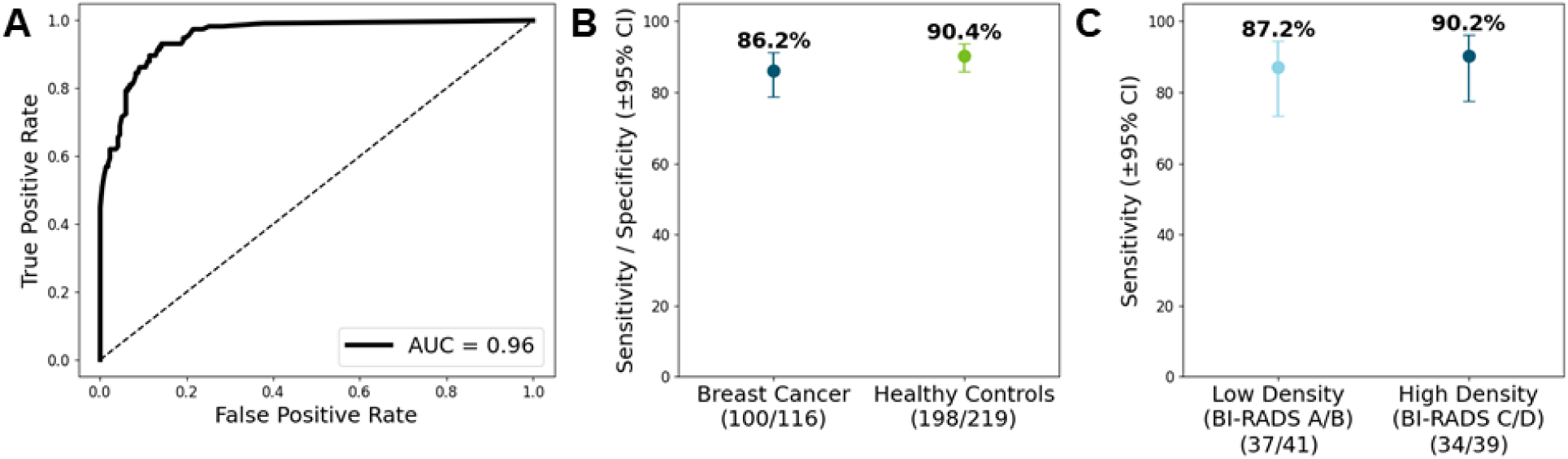
Performance of the protein-based ML classifier. (A) Receiver operating curve (ROC) showing the model’s strong discriminatory power to distinguish between breast cancer patients and healthy controls. (B) Dot plot of the overall sensitivity and specificity of the ML classifier. (C) Dot plot of the sensitivity of the ML classifier broken down by breast density. Here, low density is defined as women with BI-RADS A or B, while high density is defined as women with BI-RADS C or D. Brackets on dot plots represent the 95% Wilson confidence intervals on the measurements.

To emphasize this workflow’s potential as a diagnostic tool for women with dense breasts, we evaluated differences in the model’s accuracy based on breast density. For breast cancer patients with both low (BI-RADS A/B) and high (BI-RADS C/D) breast densities, the model maintains a sensitivity of greater than 85% across both groups (low density: 87.2% (73.3%-94.4%), high density: 90.2% (77.5%-96.1%)) at a set specificity of 90.4% (Figure 4C).

## Discussion

Liquid biopsy-based proteomics holds the potential to advance the sensitivity of early-stage cancer detection, leading to increased diagnosis in patient populations where conventional screening methods are insufficient. Despite the surge of interest in liquid biopsies over the past decade for their application in treatment monitoring and molecular residual disease (MRD) detection, their use as a diagnostic tool remains limited. Several technologies have emerged for early-stage cancer detection using a variety of blood-based cancer analytes. Exai Bio demonstrated the utility of oncRNA for the sensitive detection of early-stage lung cancer [21]. Preliminary findings from Nexosome have also shown the diagnostic potential of EV proteomic profiling for early-and late-stage multi-cancer detection [22]. While these assays show promise, there are limited results for the detection of early-stage breast cancer. Other early cancer detection tests have sought to combine multiple cancer analytes in the hopes of increasing diagnostic accuracy. The CancerSEEK assay, which combines targeted mutational analysis of ctDNA with the quantification of select protein biomarkers, showed strong results for certain cancer types, but reported poor sensitivity for breast cancer (33%) [23]. In this study, we aim to overcome these limitations by developing an accessible, proteomic-based liquid biopsy specifically for early-stage breast cancer detection.

Other high-throughput proteomic technologies have been developed to perform deep proteomic profiling. Olink’s antibody proximity extension assay (PEA) and SomaLogic’s aptamer SomaScan platform are two affinity-based proteomic assays that have been reported to identify a median of 2,941 and 4,719 proteins per patient, respectively, in recent studies [24], [25]. Seer’s Proteograph Assay also uses a MS-based approach following nanoparticle protein enrichment, and, in a recent study, reported a median of 2,899 proteins detected in >20% of 345 plasma samples [26]. Here, we captured a median of over 6,500 proteins per patient using our MS-based analysis in both cancer and non-cancer individuals. Compared to other high throughput methods, the depth of our results highlights the strength of our analysis workflow and underscores our potential to screen for low abundant proteins within the plasma proteome (Figure S3).

We designed a novel DIA analysis normalization pipeline to ensure we could process samples in a robust and computation-efficient manner. By normalizing each sample to a set of 100 “normalizing samples”, we ensured that the protein abundance quantifications were directly comparable across samples, making them ideal for a downstream machine learning pipeline. To validate the efficacy of this approach, we aimed to understand the marginal effect of adding one extra sample to the normalization set. We randomly selected four normalization subsets consisting of 10, 20, 50, and 100 samples, respectively. For each subset, one additional randomly chosen sample was added, and MaxLFQ-normalized protein group (PG) abundance values were calculated. This process was repeated three times for each normalization subset. The results demonstrated that the stability of protein quantification increases with the size of the normalization set, with minimal impact from the addition of a single sample at larger subset sizes (Figure 5). This strategy allows us to process future samples independently using the precomputed protein quantifications of the normalizing samples via MaxLFQ without having to rerun the DIA analysis on the entire dataset.

**Figure 5.**
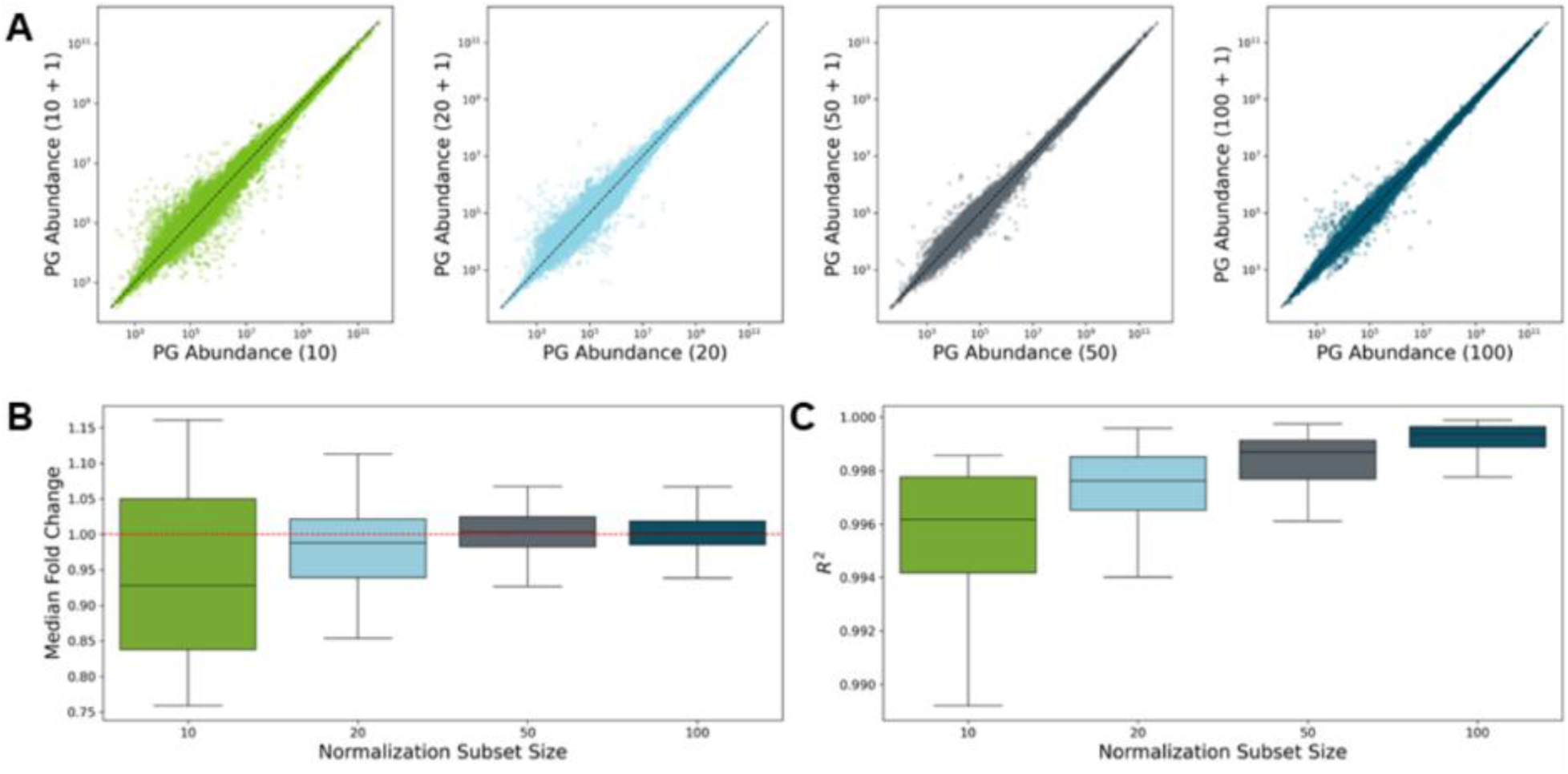
Methodology of protein normalization strategy. (A) Scatter plot of all the PG abundances normalized for n vs (n+1) runs. (B) Boxplot for median fold change by run of PG abundances normalized for n vs (n+1) runs. (C) R^2^ fit for PG abundances normalized for n vs (n+1) runs.

To encapsulate information from the full range of proteins identified in this study, we trained a logistic regression classifier with L1 regularization to detect early-stage breast cancer based on patient proteome profiles. This approach simultaneously regularizes the classifier to improve prediction accuracy while performing variable selection by shrinking the coefficients of less informative proteins to zero. Given a relatively limited sample size, we used a LOOCV approach to obtain nearly unbiased estimates of the performance metrics. Recent studies have also explored the potential of MS-based plasma proteomics in early-stage breast cancer detection [27], [28]. Unlike the workflow presented here, these studies focus primarily on the use of MS for protein biomarker discovery; they then validate the diagnostic capability of candidate biomarkers in an early-stage breast cancer cohort. While these studies report strong prediction accuracy, with reported AUC values ranging from 0.91-0.93, their sample sizes were limited (10-20 breast cancer patients per cohort). Furthermore, set protein panels may have limited diagnostic capabilities due to inherent cancer heterogeneities. The use of our ML method provides a robust, unbiased evaluation of the full scope of detected plasma proteins, potentially explaining the increased performance of our classification method.

There are a few notable limitations to this study. First, despite the classification model’s strong predictive performance, the sample size for this study is modest and lacks an independent validation cohort. Increasing the sample size of both breast cancer cases and controls will help to increase the performance of our ML classifier and precision of these results. In future work, a larger validation study will be conducted consisting of a cohort of greater than 1,000 participants to verify the current findings. Second, the cancer patient and healthy control groups were not age matched in this study. While younger healthy individuals were included to reduce the risk of adding confounding effects from false negatives to the classifier, additional validation will be required to elucidate any age-related effects on model performance. Third, the patients within the breast cancer cases were predominantly white (84.5%), potentially limiting the validity of these findings for minority groups. Finally, our current methodology requires blood to be drawn into EDTA tubes and shipped overnight for analysis. Future efforts will be aimed at reducing age and racial disparities in our validation cohorts and evaluating other ways to mitigate protein variations that come from blood stored in EDTA tubes, with the goal of increasing the accessibility and consistency of recruited samples.

## Conclusion

Deep proteomic analysis holds incredible potential as a tool for early cancer detection. We demonstrate, using a liquid biopsy-based test, how unbiased, MS-based analysis of the plasma proteome can detect a large dynamic range of protein abundances over 8 orders of magnitude in early-stage breast cancer. Our machine learning method leveraged this broad scope of data acquired for disease classification, expanding our ability to analyze complex proteome profiles while also providing a powerful diagnostic tool. While this workflow is currently limited to breast cancer detection, this pipeline can be broadly adapted to other types of cancer and diseases.

## Data Availability

The data sets analyzed in this study are available upon reasonable request by email to the corresponding author.

## Acknowledgements

We would like to thank all the brave patients who donated blood for this study as well as the clinical research staff and physicians within the Allina health system who recruited and collected samples from these patients. We would also like to thank the entire staff at Astrin Biosciences for their continued efforts to end cancer as we know it.

## Authors’ Contributions

Conceptualization: **AH**, **JMD**; Resources: **SP**, **BRK**, **YO**; Data curation: **AH**, **YT**, **KM**, **KJK**, **NRB**, **SYB**; Formal analysis: **AH**, **YT**, **KM**, **NRB**; Supervision: **CGG**, **JMD**; Validation: **JMD**; Investigation: **AH**, **GS**, **JMD**; Visualization: **YT**, **KJK**, **HB**, **JMD**; Methodology: **AH**, **GS**, **SYB**, **JMD**; Writing – original draft: **AH**, **YT**, **KJK**, **HB**, **JMD**; Project administration: **KJK**, **CGG**, **ARH**, **JMD**; Writing – review and editing: **AH**, **YT**, **KM**, **GS**, **KJK**, **NRB**, **CGG**, **SYB**, **HB**, **ARH**, **SP**, **BRK**, **YO**, **JMD**.

